# Efficacy of vasopressin, steroid, and epinephrine protocol for in-hospital cardiac arrest resuscitation: A systematic review and meta-analysis of randomized controlled trials with trial sequential analysis

**DOI:** 10.1101/2021.11.20.21266625

**Authors:** Danish Iltaf Satti, Yan Hiu Athena Lee, Keith Sai Kit Leung, Jeremy Man Ho Hui, Thompson Ka Ming Kot, Arslan Babar, Abraham KC Wai, Tong Liu, Leonardo Roever, Gary Tse, Jeffrey Shi Kai Chan, International Health Informatics Study (IHIS) Network

## Abstract

**Aim:** To assess the effect of vasopressin, steroid and epinephrine (VSE) combination therapy on return of spontaneous circulation (ROSC) after in-hospital cardiac arrest (IHCA), and test the conclusiveness of evidence using trial sequential analysis (TSA).

**Methods:** The systematic search included PubMed, EMBASE, Scopus, and Cochrane Central Register of Controlled Trials. Randomized controlled trials that included adult patients with in-hospital cardiac arrest, with at least one group receiving combined vasopressin, epinephrine and steroid therapy were selected. Data was extracted independently by two reviewers. The main outcome of interest was ROSC. Other outcomes included survival to hospital discharge with good neurological outcomes and survival to 30 and 90 days with good neurological outcomes.

**Results:** We included a total of three randomized controlled trials (n=869 patients). Results showed that Vasopressin, steroid and epinephrine combination therapy increased return of spontaneous circulation (risk ratio, 1.32; 95% CI, 1.18-1.47) as compared to placebo. Trial sequential analysis demonstrated that the existing evidence is conclusive. This was also validated by the alpha-spending adjusted relative risk (1.32 [1.16, 1.49], p<0.0001). Other outcomes could not be meta-analysed due to differences in timeframe in the included studies.

**Conclusions:** VSE combination therapy administered in cardiopulmonary resuscitation led to improved rates of return of spontaneous circulation. Future trials of vasopressin, steroid and epinephrine combination therapy should evaluate survival to hospital discharge, neurological function and long-term survival.

## Introduction

Cardiac arrest is the sudden cessation of cardiac activity. Most randomized controlled trials (RCTs) on cardiac arrest pharmacotherapy focused on out-of-hospital cardiac arrest, while in-hospital cardiac arrest (IHCA) received less attention.^1^ While epinephrine has been the mainstay for managing cardiac arrest, it may cause myocardial ischaemia and dysfunction.^2^ This is circumvented by vasopressin, which causes systemic vasoconstriction with sparing of coronary or cerebral vasculature.^3^ Furthermore, corticosteroid has important vascular effects which make it potentially beneficial in treating cardiac arrest.^4^ Therefore, combined vasopressin, steroids and epinephrine therapy (VSE) in IHCA has been actively investigated.

Nonetheless, the value of VSE remained controversial, and conclusiveness of the underlying evidence was unclear. We thus performed this systematic review and meta-analysis to evaluate the efficacy of VSE in IHCA, with trial sequential analysis (TSA) for assessing the conclusiveness of existing evidence.

## Materials and methods

This systematic review and meta-analysis was conducted according to the preferred reporting items for systematic reviews and meta-analyses (PRISMA) statement and the Cochrane Handbook for Systematic Reviews of Intervention (PROSPERO registration: CRD42021284320). Ethics approvals were not required for this study as it is a systematic review and meta-analysis.

### Search strategy and selection criteria

PubMed, Scopus, and EMBASE were searched electronically for all studies concerning cardiac arrest resuscitation regardless of publication type or language. All databases were searched from inception till 4th October 2021. The search string was (“Heart Arrest” OR “Cardiac Arrest” OR “Asystole” OR “Cardiopulmonary Arrest” OR “Ventricular Tachycardia” OR “Ventricular Fibrillation” OR “Pulseless electrical activity” OR “Cardiopulmonary Resuscitation” OR “Advanced Cardiac Life Support”) AND (“Vasopressin” OR “Antidiuretic Hormones”) AND (“Epinephrine” OR “Adrenaline”) AND (“Steroids” OR “Methylprednisolone” OR (“Glucocorticoids” OR “Corticosteroids” OR “Corticoids”) OR “Hydrocortisone” OR “Dexamethasone”). References of relevant papers were manually searched for eligible articles. We also searched ClinicalTrials.gov and the Cochrane Central Register of Controlled Trials (2003 to present) for ongoing, incomplete, or unpublished studies.

All randomized controlled trials (RCT) that studied adult patients with IHCA, with ≥1 group receiving VSE, were included. Only studies written in English were included. Single-arm studies, conference publications, case reports/series, observational studies, and reviews were excluded.

### Risk of bias assessment

Cochrane Risk of Bias 2 tool was used by two independent reviewers (DIS, JMHH) for risk of bias assessment. Conflicts were settled by an independent reviewer (JSKC).

### Data extraction and outcomes

Two reviewers (DIS, YHAL) independently extracted data from included studies, including country, sample sizes, patients’ demographics, IHCA rhythm, intervention type, VSE dosage, intervention timing, and outcomes. Conflicts were resolved by an independent reviewer (JSKC). The primary outcome was sustained return of spontaneous circulation (ROSC) as defined in the included studies.

The secondary outcomes, as reported by the included trials, were also qualitatively described and compared. Nonetheless, due to the differences in timeframe used for the secondary outcomes in the included trials, meta-analysis of these outcomes was not possible and thus was not performed.

### Statistical Analysis

Review Manager (v5.4, The Cochrane Collaboration, 2020) was used for statistical analysis. Relative risk (RR) with 95% confidence interval (CI) was used as the summary statistic. Random effects model was used with the Mantel-Haenszel test. Heterogeneity was assessed by Chi^2^-test and I^2^; I^2^≥0.40 was considered to imply significant heterogeneity.

TSA was performed using a combination of sample and event sizes. TSA controls and adjusts for type-1 error, minimizing the risks of spuriously positive results.^5^ The O’Brien-Fleming α-spending function was used to adjust the Z-score threshold. The required information size was estimated using a ±20% *a priori* relative risk reduction, which commonly represents clinically significant and realistic effect sizes. Control arm incidence was calculated from included patients. Permissible two-sided 5% type-1 error, and 20% type-2 error were used. TSA was performed using Copenhagen Trial Unit TSA software v0.9.5.10beta.

## Results

The literature search is summarized in **Figure 1**: 392 non-duplicate citations were identified; after full-text screening of six, three were included with 869 patients in total (VSE: 415, control: 454). Electronic search of Clinicaltrials.gov and the Cochrane Central Register of Controlled Trials identified no relevant ongoing, incomplete, or unpublished study.

**Figure 1.**
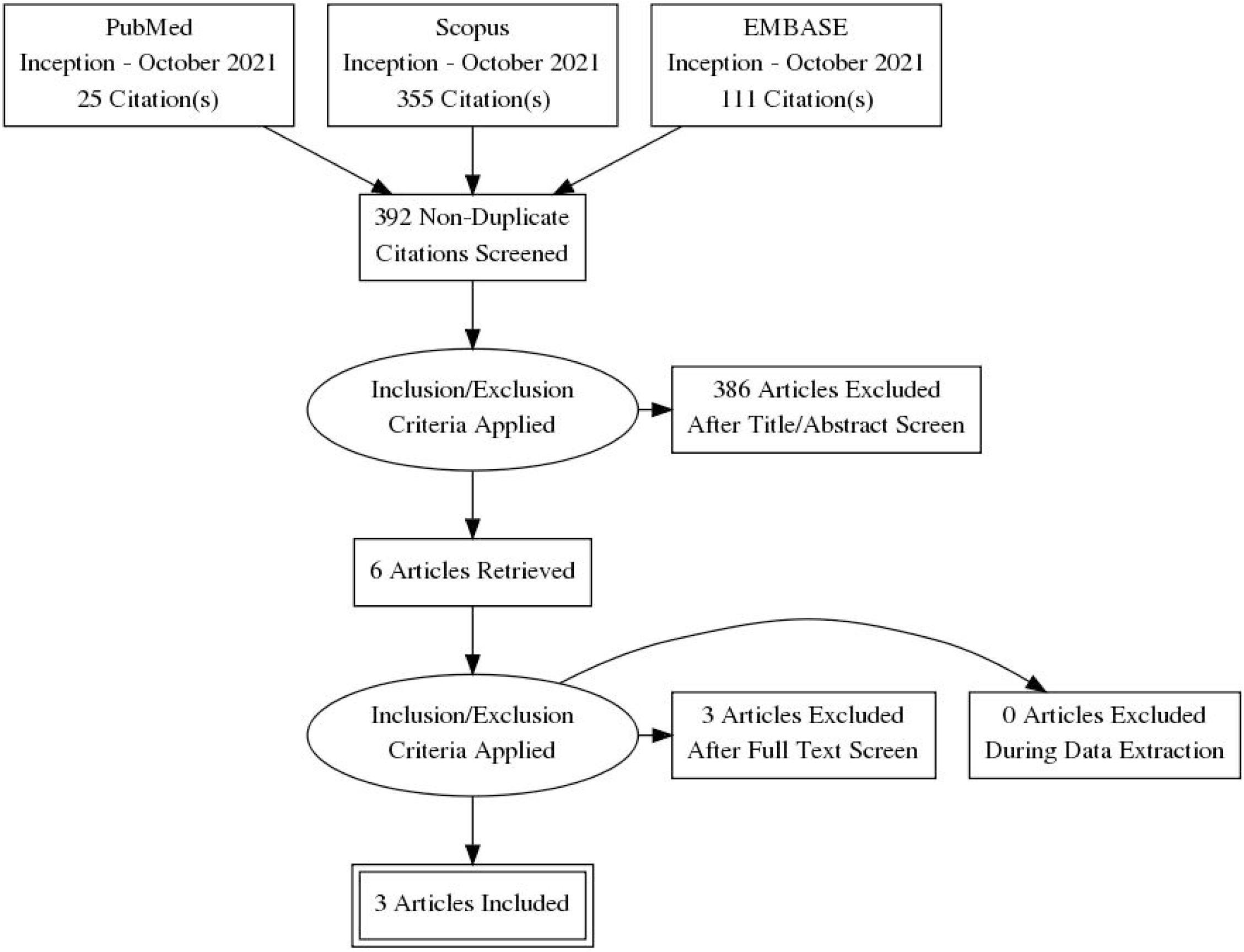
PRISMA flow diagram detailing the search strategy and study selection process for this systematic review and meta-analysis on the efficacy of vasopressin-steroid-epinephrine combination therapy for in-hospital cardiac arrest resuscitation

### Quality assessment

Risk of bias of the included studies were summarized in a **Figure 2**. Two had low risk of bias.^6,7^ There were some concerns for one due to lacking an identifiable pre-specified analysis plan.^8^ However, the results were unlikely to be selected from multiple outcome measurements or analyses. The effects of such concerns were thus deemed minimal, and the study was included in the final meta-analysis.

**Figure 2.**
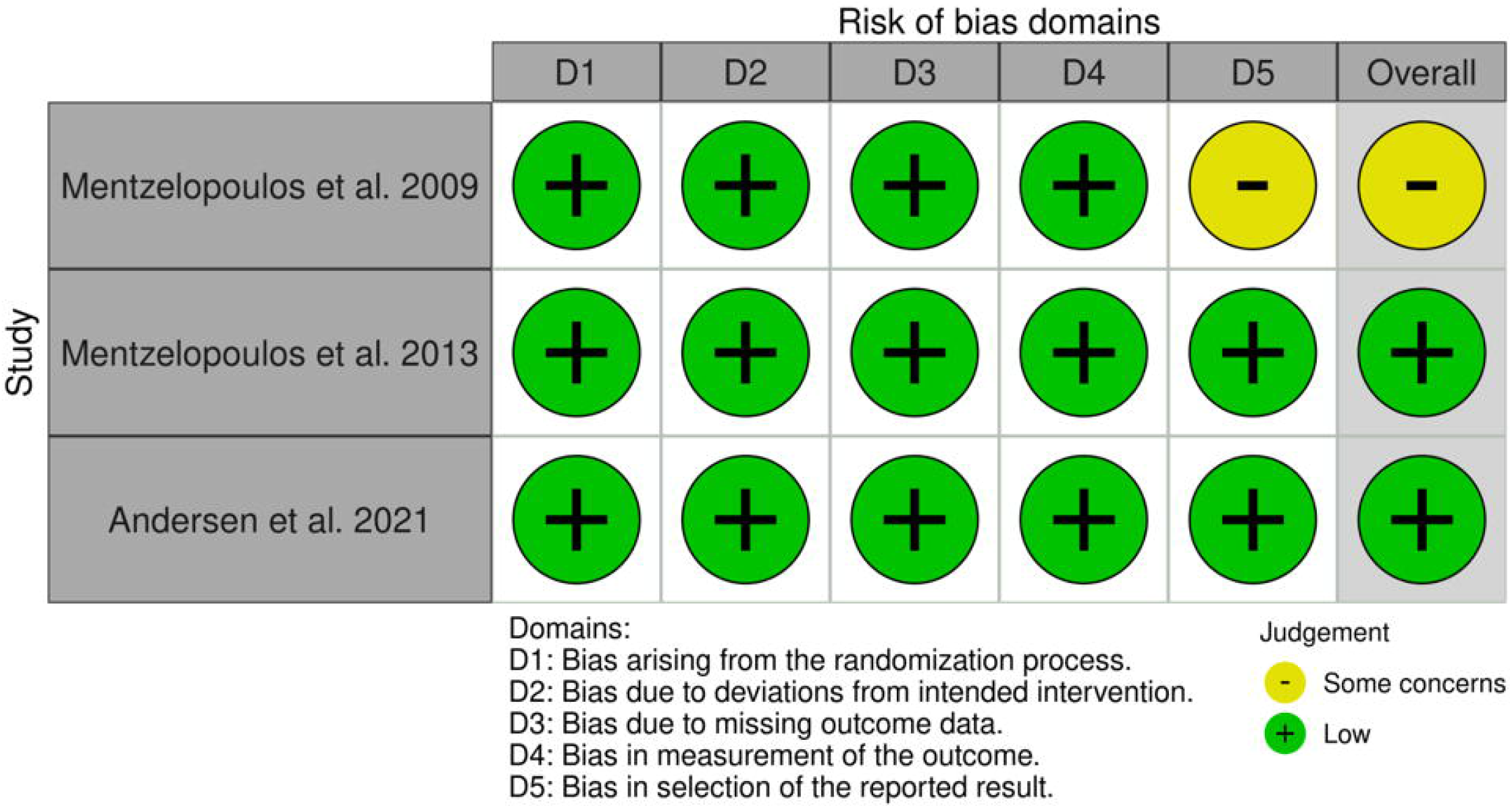
Risk of bias (quality) assessment of randomized controlled trials using Cochrane Risk of Bias 2 tool

### Baseline characteristics

The baseline characteristics for included studies were summarized in a **Supplementary Table 1**. VSE was used during and after IHCA in two ^6,7^, while only during IHCA in the third.^8^ Patients were followed up for 60 days in the former two ^6,7^, and 30 and 90 days in the latter.^8^

### Primary outcome (sustained ROSC)

Sustained ROSC was defined, by the included studies, as recovery of circulation alongside reestablishment of blood pressure and pulse for ≥15 (7) or ≥20 minutes ^6,8^. Meta-analysis showed significantly higher rates of sustained ROSC with VSE (RR 1.32 [1.18-1.47], p<0.00001; **Figure 3**) without significant heterogeneity (I^2^=0%; Chi^2^=1.10, df=2, p=0.58).

**Figure 3.**
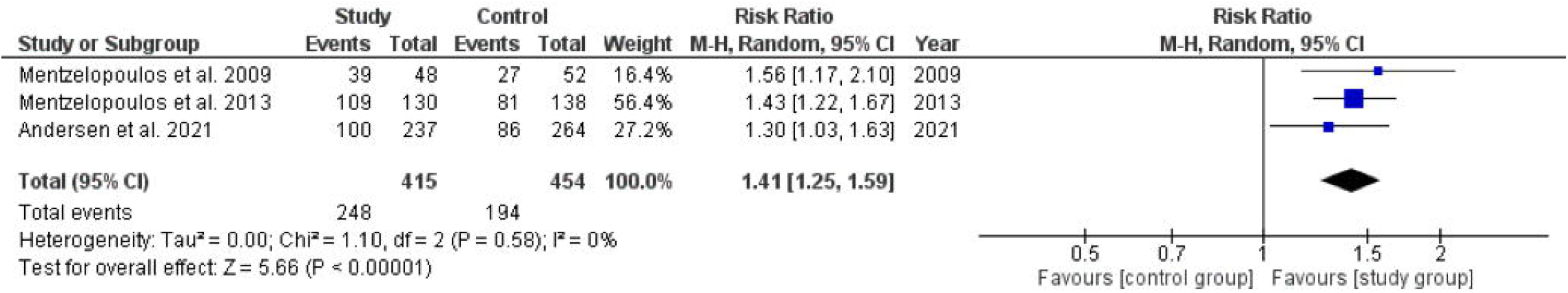
Risk ratios of randomized controlled trials comparing vasopressin-steroid-epinephrine combination therapy to placebo using a random-effects model with Mantel-Haenszel (M-H) weighting for return of spontaneous circulation (ROSC).

TSA results are presented graphically (**Figure 4**). The blue line (Z-curve) represents the cumulative effect size, with individual trial effects displayed chronologically. The red line represents the TSA-adjusted significance boundary. The dotted green line represents the conventional, unadjusted significance boundary with 2-sided 5% type-1 error. Figure 4 showed the Z-curve crossing both adjusted and unadjusted significance boundaries, implying that the existing evidence is sufficient to conclusively demonstrate a 20% increase in likelihood of sustained ROSC in the VSE group. Thus, VSE conclusively increased the likelihood of sustained ROSC in IHCA, as further corroborated by α-spending-adjusted RR (1.32 [1.16-1.49], p<0.0001).

**Figure 4.**
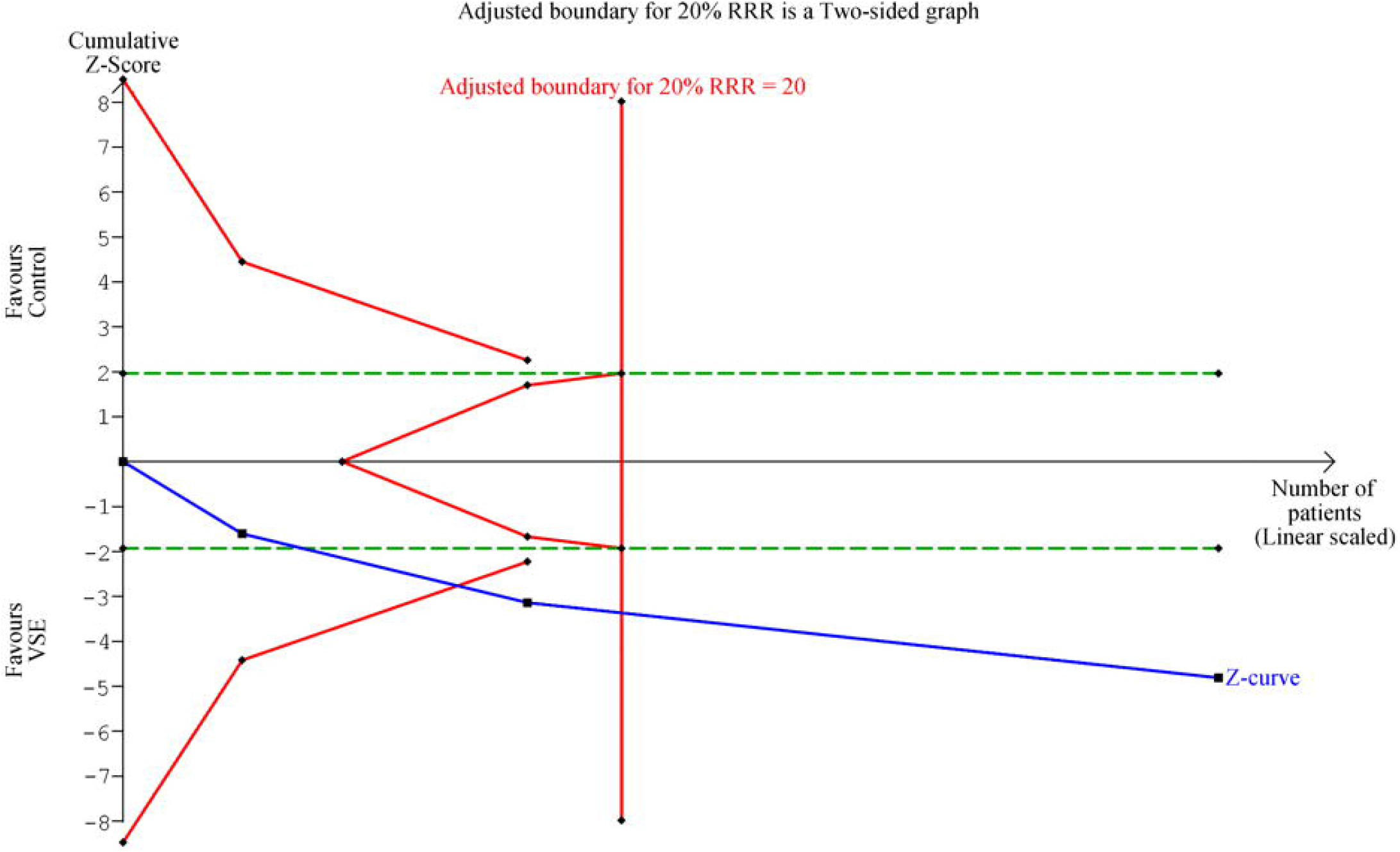
Result from trial sequential analysis. The existing evidence supporting the use of the vasopressin-steroid-epinephrine combination therapy in in-hospital cardiac arrest in terms of sustained return of spontaneous circulation is conclusive.

### Other outcomes as specified by the included studies

Besides sustained ROSC, the included studies also reported survival to hospital discharge with good neurological outcome ^6,7^, and survival to 30 and 90 days with good neurological outcomes.^8^

In both studies by Mentzelopoulos *et al*. ^6,7^, survival was measured till hospital discharge, while Anderson *et al*. measured survival at 30 and 90 days.^8^ While both studies by Mentzelopoulos *et al* found significant survival benefits with VSE, Anderson *et al* did not observe any at both 30 and 90 days.

Both studies by Mentzelopoulos *et al* reported neurological status at the time of hospital discharge using the Glasgow-Pittsburgh Cerebral Performance Category (CPC) ^6,7^, while Anderson *et al* (8) used the modified Rankin scale alongside Glasgow-Pittsburgh CPC score to assess neurological outcomes at 30 and 90 days. While the former two found neurological benefits with VSE, Anderson *et al* did not observe any.

### Adverse events

There were no adverse events reported in both studies by Mentzelopoulos *et al* ^6,7^, while Anderson *et al* reported hypernatremia and hyperglycaemia in patients who achieved ROSC.^8^ However, the rate of adverse events in both the study and control group were nearly similar. Hyperglycaemia occurred in 77% and 73% of patients in study and control group respectively. Similarly, hypernatremia occurred in 28 % and 31 % in the study and control group respectively.

## Discussion

This meta-analysis found that VSE was associated with significantly improved rate of sustained ROSC in IHCA. Trial sequential analysis further demonstrated that this could be considered conclusive.

Shah *et al*.^9^ and Liu *et al*.^10^ previously meta-analysed the efficacy of steroid therapy for cardiac arrest, both including VSE in subgroup analyses. Our findings agree with theirs. Nonetheless, to the best of our knowledge, this was the first meta-analysis using TSA to evaluate VSE. TSA builds on the fact that the conclusiveness of any evidence increases with the cumulative sample size, and larger effects require fewer subjects for observations to be conclusive.^5^ When meta-analysing high-quality trials without substantial risks of bias, as is the case in this meta-analysis, TSA is appropriate for demonstrating conclusiveness of observed meta-analytical effects.^5,11^ Our results implied that further studies are not required to evaluate VSE in IHCA in terms of sustained ROSC; survival and neurological outcomes should be the focus instead.

VSE may be beneficial via several mechanisms. Persistent β_2_-receptor activation by epinephrine predisposes to post-resuscitation myocardial dysfunction^2^ – contrastingly, vasopressin has no direct myocardial effect, and dilates cerebral vessels more effectively which avoids myocardial ischaemia.^3^ Meanwhile, benefits of corticosteroid may be attributed to its role in maintaining myocardial contractility, endothelial integrity, vascular tone, and vascular responsiveness to catecholamine.^4^

By using TSA, we confirmed the certainty of our findings i.e. VSE is associated with improved likelihood of sustained ROSC, which is the main goal of CPR. Although the long-term outcomes are unclear, current evidence suggests that VSE is safe with very low rates of adverse events reported, hence its benefits outweigh the risks. Given the grave prognosis of cardiac arrest with less than 12% surviving ^12^, and the extremely limited range of available therapies, it is reasonable to start using VSE in clinical management of IHCA, while conducting future studies to further delineate its long-term effects simultaneously.

### Limitations

This meta-analysis has several limitations. First, subgroup analysis by the rhythm of IHCA was impossible since none of the included studies reported rhythm-specific findings. Second, survival and neurological outcomes could not be meta-analysed due to the different timeframes reported in the included studies. Therefore, despite TSA showing that the current evidence for sustained ROSC is conclusive, further studies focusing on the effects of VSE on survival and neurological outcomes are still needed. Third, with only three studies, publication bias could not be assessed. Nonetheless, no incomplete or unpublished trials were identified by electronic search of Clinicaltrials.gov and the Cochrane Central Register of Controlled Trials, suggesting that likelihood of significant publication bias is low.

## Conclusions

This meta-analysis of RCTs demonstrated conclusively that VSE led to improved rates of ROSC. Future trials should assess other outcomes of VSE, such as survival, neurological recovery, and longer-term outcomes.

## Data Availability

All data produced in the present work are contained in the manuscript.

## Abbreviations^1^

^1^ VSE: Vasopressin, steroid and epinephrine
ROSC: Return of spontaneous circulation
TSA: Trial sequential analysis
IHCA: In-hospital cardiac arrest

## Legends to figures

**Supplementary Table 1**. Baseline characteristics of the included studies

