## Supplementary Table 1 for "Efficacy of vasopressin, steroid, and epinephrine protocol for in-hospital cardiac arrest resuscitation: A systematic review and meta-analysis of randomized controlled trials with trial sequential analysis"

**Supplementary Table 1.** Baseline characteristics of included studies

| Author, year | Total Sample Size | Country | Time of intervention | Groups | Type of therapy | Age, mean (SD), year | Males, N (%) | Drug dosage | Rhythm of Cardiac Arrest | Outcomes | Follow Up |
|---|---|---|---|---|---|---|---|---|---|---|---|
| Mentzelopoulos et al. 2009[7] | 100 | Greece | During and after resuscitation | Study Group (n=48) | Vasopressin<br>Epinephrine<br>Methylprednisolone<br>Hydrocortisone for Post resuscitation shock | 65.4 (17.6) | 30 (63%) | Vasopressin, IU, mean (SD)= 73.3 (30.1)<br><br>Epinephrine, IU, mean (SD)= 6.3 (5.8)<br><br>Methylprednisolone, mg, mean (SD)= 40.0 (0.0)<br><br>Hydrocortisone (300mg daily for 7 days maximum and gradual taper) | Asystole=63%<br><br>Ventricular fibrillation/tachycardia=15%<br><br>Pulseless electrical activity=23% | Primary=ROSC, Survival to Hospital Discharge with good neurological outcomes | 60 days |
| | | | | Control Group (n=52) | Saline placebo<br>Epinephrine | 69.2 (17.7) | 29 (56%) | Epinephrine, mg, mean (SD)= 7.8 (7.0) | Asystole=60%<br><br>Ventricular fibrillation/tachycardia=13%<br><br>Pulseless electrical activity=27% | | |
| Mentzelopoulos et al. 2013[6] | 268 | Greece | During and after resuscitation | Study Group (n=130) | Vasopressin<br>Epinephrine<br>Methylprednisolone<br>Hydrocortisone for Post resuscitation shock | 63.2 (17.6) | 95 (73.1%) | Vasopressin, IU, mean (SD)= 70.3 (31.2)<br><br>Methylprednisolone, mg, mean (SD)=40.0 (0.0)<br><br>Epinephrine, mg, median (IQR)=4(2-5)<br><br>Hydrocortisone (300mg daily for 7 days maximum and gradual taper) | Asystole=63.8%<br><br>Ventricular fibrillation/ tachycardia=16.9%<br><br>Pulseless electrical activity=19.2% | Primary=ROSC, Survival to Hospital Discharge with good neurological outcomes | 60 days |

| | | | | | | | | | | | |
|---|---|---|---|---|---|---|---|---|---|---|---|
| | | | | Control Group (n=138) | Saline placebo<br>Epinephrine | 62.8 (18.6) | 88 (63.8%) | Epinephrine, mg, median (IQR)=5(3-9) | Asystole=70.3%<br><br>Ventricular fibrillation/ tachycardia=16.7%<br><br>Pulseless electrical activity=13% | | |
| Andersen et al. 2021[8] | 512 | Denmark | During resuscitation | Study Group (n=237) | Vasopressin<br>Epinephrine<br>Methylprednisolone | 71 (13) | 148 (62%) | Vasopressin, IU=20 per dose (maximum 4 doses)<br><br>Methylprednisolone, mg= 40 per dose<br><br>Epinephrine= Not mentioned | Asystole=35%<br><br>Ventricular fibrillation/ tachycardia=7%/2%<br><br>Pulseless electrical activity=57% | Primary=ROSC<br><br>Secondary=Survival to 30 days and 30 days with good neurological outcomes | 90 days |
| | | | | Control Group (n=264) | Saline placebo<br>Epinephrine | 70 (12) | 174 (66%) | Epinephrine= Not mentioned | Asystole=36%<br><br>Ventricular fibrillation/ tachycardia=8%/3%<br><br>Pulseless electrical activity=52% | | |
